# “Susceptibility and risk of suffering SARS-COV-2 infection by demographic characteristics and pre-existing medical conditions among middle-aged and older adults in Tarragona, Spain: results from the *COVID19 TARRACO* Cohort Study, March-June 2020”

**DOI:** 10.1101/2021.02.09.21251398

**Authors:** EM. Satué-Gracia, A. Vila-Córcoles, C. de Diego-Cabanes, A. Vila-Rovira, C. Torrente-Fraga, F. Gómez-Bertomeu, I. Hospital-Guardiola, O. Ochoa-Gondar, F. Martín-Luján

**Affiliations:** Primary Care Department Camp de Tarragona. Institut Català de la Salut (ICS), Tarragona, Spain; Fundació Institut Universitari per a la recerca a l’Atenció Primària de Salut Jordi Gol i Gurina (IDIAPJGol), Barcelona, Spain; Technology and Informatic Department, Institut Català de la Salut (ICS), Tarragona, Spain; Microbiology Department. Hospital Universitari Joan XXIII. Institut Català de la Salut (ICS), Tarragona, Spain

**Keywords:** Coronavirus, SARS-COV-2, COVID19, Incidence, Risk

## Abstract

**Objective:** To analyse susceptibility/risk of suffering COVID19 among adults with distinct underlying medical conditions.

**Methods:** Cohort study (population-based) including 79,083 people ≥50 years-old in Tarragona (Southern Catalonia, Spain). At study start (01/03/2020) baseline cohort characteristics (demographic, previous comorbidities, chronic medications and vaccinations’ history) were recorded. Primary outcome was laboratory-confirmed COVID19 incurred in cohort members throughout 01/03/2020-30/06/2020. Risk of suffering COVID19 was evaluated by Cox regression, estimating multivariable hazard ratios (HRs) adjusted for age/sex and previous comorbidities.

**Results:** Across study period, 536 laboratory-confirmed COVID19 cases were observed (mean incidence: 39.5 cases per 100,000 persons-week). In multivariable-analysis, age/years (HR: 1.01; 95% CI: 1.00-1.02; p=0.050), nursing-home (HR: 20.19; 95% CI: 15.98-25.51; p<0.001), neurological disease (HR: 1.35; 95% CI: 1.03-1.77; p=0.029), taking diuretics (HR: 1.39; 95% CI: 1.10-1.75; p=0.006), antiplatelet (HR: 1.36; 95% CI: 1.05-1.76; p=0.021) and benzodiazepines (HR: 1.24; 95% CI:1.00-1.53; p=0.047) significantly increased risk; while smoking (HR: 0.57; 95%CI: 0.41-0.80; p=0.001), angiotensin converting enzyme inhibitors (HR: 0.78; 95% CI: 0.61-1.00; p=0.048), angiotensin II receptor blockers (HR: 0.70; 95%CI: 0.51-0.96; p=0.027) and statins (HR: 0.75; 95% CI: 0.58-0.96; p=0.025) were associated with reduced risk. Among non-institutionalised persons, cancer, renal and cardiac disease appeared also related to increased risk, whereas influenza vaccination was associated with reduced risk.

**Conclusion:** In a setting with relatively low incidence of COVID19 across the first wave of pandemic period, age, nursing-home residence and multiple comorbidities appear predisposing for COVID19 among middle-aged/older adults. Conversely, statins, angiotensin-receptor blockers/inhibitors and influenza vaccination were related with decreased risk.

## INTRODUCTION

The novel severe acute respiratory syndrome coronavirus 2 (SARS-COV-2), discovered in December 2019 following an outbreak of pneumonias in Wuhan, China, causes an infectious illness known as coronavirus disease 2019 (COVID19). The World Health Organisation (WHO), due to the alarming levels of spread of the disease and its severity, declared it to be a pandemic on March 11, 2020. [1] Currently the disease continues to spread in the world, with almost 100 million confirmed cases in later January 2021. [2]

In Spain there was a first wave in March-April that was considered contained on May 11, with a total of just over 250,000 diagnosed cases; the contagion curve grew again in June until reaching the maximum peak of second wave in mid-October and as of later January 2021, in the middle of the third wave, the Spanish Ministry of Health reports 2,705,001 cases and almost 58,000 deaths. [3]

There is much information updating COVID19 surveillance epidemiological data (number of cases and deaths by demographic characteristics, accumulated incidences, ratios for positive laboratory testing, etc.); but there is few population-based data on specific incidence and susceptibility for SARS-COV-2 infection in population subgroups with distinct underlying -believed as at-risk-conditions. [2,3] Earlier studies regarding clinical characteristics and prevalence of comorbidities among COVID19 patients reported that cardiovascular disease (including hypertension), chronic respiratory disease, renal disease, cancer, diabetes mellitus, obesity and smoking were major risk factors for severe COVID19. However, most of these results (mainly based on hospitalised severe cases) are likely confounded by age and other unadjusted variables (including else socio-demographic, medical or health care-related factors). [4-7] Of note, COVID19 patients managed as outpatients were generally poorly represented in COVID19 research studies. [4-7] In this sense, a UK report based on a primary care cohort found that positive SARS-COV-2 testing was associated with similar risk factors (except smoking) as described for severe COVID19 in hospital settings. [8] Nevertheless, there is uncertainty about the possibility that some of these conditions, although they increase the risk of severe COVID19, may not “per se” increase the risk of SARS-COV-2 infection. [9]

In this context, we designed a large population-based retrospective-prospective cohort study (initiated in April 2020 and named as *TARRACO Cohort Study*) aimed to investigate the incidence, clinical characteristics, susceptibility and risk profile of suffering COVID19 among middle-aged and older adults (who supported the greatest burden of COVID19 at this time) in the region of Tarragona across pandemic period. First-time analyses reporting earlier data have been published. [10,11] The present study updates population-based incidence data and risk of suffering laboratory-confirmed COVID19 among the 79,083 cohort members by distinct underlying medical conditions (previous comorbidities, vaccinations’ history and common chronic medications’ use), reporting final data observed at the end of the planned study period (from 1st March to 30th June, 2020).

## METHODS

### Design, setting and study population

The design, setting and study population have been extensively described elsewhere. [10,11] Briefly, this is a retrospective-prospective population-based cohort study (began in April 2020) that included 79,083 persons ≥50 years old in the region of Tarragona, whose total population (all-age) is 210,672 inhabitants. The cohort incorporates all persons ≥50 years old assigned to 12 Primary Care Centres (PCCs) managed by the *Institut Català de la Salut (ICS)* in the study area (*Tarragonés, Alt Camp and Conca Barberà* counties). The reference Hospital and Laboratory for the 12 participating PCCs were *Hospital Universitari Joan XXIII* and its Microbiological Department in the city of Tarragona. According to census data, the study cohort represents approximately 75% of inhabitants aged 50 years or older in the area. [12] Cohort members were followed since March 1, 2020 (date before the onset of the epidemic in the region), until the appearance of any study event (COVID19 diagnosis or death), or until the end of 4-month follow-up (June 30, 2020).

The study was approved by the Ethics Committee of the Foundation University Institute for Primary Health Care Research (*IDIAP Jordi Gol*, Barcelona, file 20/065-PCV) and was conducted according to the Helsinki Declaration and Spanish legislation on biomedical studies, data protection and respect for human rights. [13] Considering public health surveillance emergency, informed consent for all cohort members was waived.

### Data sources

An institutional clinical research database earlier used for other cohort studies in the area (CAPAMIS), [14] was updated and became the primary data source for this COVID19 epidemiologic report; it includes administrative and clinical data (coded according to the International Classification of Diseases 10th Revision, ICD-10), extracted from the PCCs’ clinical records electronic system, that operates since the 2000s. We collected socio-demographic characteristics, previous comorbidities,chronic medication and vaccinations’ history for cohort members in order to establish their baseline conditions (at March 1, 2020).

At the COVID19 epidemic period onset in the area, two electronic alerts, linked to COVID19 laboratory tests results and ICD-10 codes for disease suspicion (B34.2 and B97.29), were added to the electronic clinical records system of PCCs. Subsequently, both data sources were associated in the anonymised research database used for this report. Vital status for all cohort members was considered according to administrative data periodically updated in the electronic PCCs’ clinical records.

### Outcomes

The main outcome was laboratory-confirmed COVID19 diagnosed by reverse transcription-polymerase chain reaction (RT-PCR) or serological testing. For laboratory diagnosis of COVID19, guidelines of the Health Department of the *Generalitat de Catalunya* were implemented. [15] Of note, the availability of PCR testing was scarce during the first weeks of epidemic period, and they were prioritised for severe cases (requiring hospital admission) and nursing-home residences (where several outbreaks took place); while less PCR tests were made among suspected cases with mild symptoms managed as outpatients. We report, in descriptive analysis, presumed COVID19 cases (people who presented clinical suspicion who were no PCR-tested) and, also, COVID19-excluded cases (with negative result in PCR-test) detected among cohort members across the study period.

### Covariates

Besides socio-demographic (age, sex and type of residence as community-dwelling/nursing-home), main covariates for this study were vaccinations’ history (influenza vaccine in prior autumn, pneumococcal [pneumococcal polysaccharide vaccine -PPV23- and pneumococcal conjugate vaccine -PCV13] or tetanus vaccination at any time) and presence of pre-existing comorbidities. The following comorbidities/underlying conditions were considered according to ICD-10 codes data registered in the electronic PCCs clinical records on 01/03/2020: neurological disease (including dementia and stroke), cancer (solid organ or haematological neoplasia (diagnosed in past 5 years), chronic renal failure, systemic autoimmune rheumatic diseases (including rheumatoid arthritis and lupus), chronic respiratory disease (including chronic bronchitis/emphysema and/or asthma), chronic heart disease (including congestive heart failure, coronary artery disease and other cardiopathies), atrial fibrillation, chronic liver disease (including chronic hepatitis and cirrhosis), arterial hypertension, diabetes mellitus, hypercholesterolemia, obesity, alcoholism and smoking.

In addition, baseline use of some common chronic medications, which could be hypothetically related with physiopathologic mechanism of SARS-COV-2 infection or virulence (e.g., antihypertensive, antiplatelet/anticoagulant and/or anti-inflammatory drugs), were also considered as exposure co-variables possibly related with the occurrence of COVID19 for the present study. Thus, active medication treatments in each cohort member on 01/03/2020, coded according to the Anatomical, Therapeutic, and Chemical classification system (ATC) of the World Health Organization, [16] were identified from the patient treatment plan registered in the PCC’s clinical records system, including the following therapeutic groups: antihypertensive (diuretics, beta-blockers, angiotensin-converting enzyme inhibitors [ACEIs], angiotensin II receptor blockers [ARBs], calcium channel blockers), statins, anticoagulants (warfarin and new oral anticoagulant drugs), antiplatelet drugs, anti-diabetic drugs (insulin, oral anti-diabetic drugs), inhaled respiratory drugs, antineoplastic agents, systemic corticosteroids, non-steroidal anti-inflammatory drugs (NSAIDs), chloroquine/hydroxychloroquine, antihistamines, proton-pump inhibitors and benzodiazepines, were registered. Criteria used to define and collect pre-existing conditions and chronic medications use are extensively described in an appendix elsewhere. [11]

### Statistical analyses

We calculated frequencies and percentages, means and standard deviations (SD) for qualitative and continuous variables, respectively. Incidence rates (IRs) for laboratory-confirmed COVID19 cases were calculated for persons-weeks, considering in the denominator the sum of time contributed by each cohort member during the study period (17.4 weeks). Confidence intervals (CIs) for IRs were calculated assuming a Poisson distribution for uncommon events. In bivariate analyses, baseline characteristics according to final presence/absence of study outcome (COVID19 diagnosis) were compared using Chi-squared or Fisher’s test as appropriate.

Cox regression analyses were used to calculate unadjusted, age&sex-adjusted and multivariable-adjusted hazard ratios (HRs) aimed at estimating the association between baseline exposure conditions and the time to the first outcome (laboratory-confirmed COVID19) in cohort members throughout the study period (from 1st March to 30th June 2020). We included in exploratory multivariable Cox models all above mentioned exposure and covariates (i.e., age, sex, residence, vaccinations’ history, comorbidities/underlying conditions and medication use). The method to select a subset of variables to include in final models was the purposeful selection. [17] These models incorporate significant and confounder variables as well as all covariates judged clinically or epidemiologically relevant. We performed a main analysis with the total study cohort (N=79,083) and two subgroup analyses restricted to community-dwelling people (N=77,669) and nursing-home residents (N=1414). Statistical significance was set at p <0.05 (two-tailed). Analyses were performed using IBM SPSS Statistics for Windows, version 24 (IBM Corp., Armonk, N.Y., USA).

## RESULTS

Study cohort included 37,626 (47.6%) men and 41,457 (52.4%) women, with a mean age of 65.8 (SD: 11.3) years. Stratifying by age, 42,684 (54%) were 50-64 years and 10,386 (13.1%) were 80 years or more; by residence, 1414 (1.8%) were nursing-home residents and 77,669 (98.2%) community-dwelling individuals.

Total cohort members were followed for 1,356,358 persons-weeks (1,335,069 persons-weeks community-dwelling and 21,289 persons-weeks nursing-home residents).

Of the 79,083 cohort members, 4084 (5.2%) were PCR tested for SARS-COV-2 infection across the study. Of them, 3577 (87.6%) had a negative result and 507 (12.4%) had a positive result (319 community-dwelling and 188 nursing-home residents). Additionally, 29 cohort members had a positive laboratory serological testing for SARS-COV-2 (twenty-six community-dwelling and three nursing-home residents). In two hundred and eighty-three cohort members COVID19 was suspected (clinically compatible) but no laboratory test was performed.

Of the 536 laboratory-confirmed COVID19 cases, 235 (43.8%) were men and 301 (56.2%) women, with a mean age of 74 years (SD: 14.4). By age groups, 177 (33.0%) occurred in those who were 50-64 years, 137 (25.6%) in those 65-79 years and 222 (41.4%) in aged 80 years or more. Three hundred and forty-five cases (64.4%) occurred in community-dwelling and 191 (35.6%) in nursing-home residents. The most prevalent comorbidities were hypertension (54.1%), cardiac disease (28.5%), obesity (25.2%), diabetes (23.5%), respiratory disease (14.9%) and neurological disease (14.7%).

Figure 1 shows weekly distribution of COVID19-confirmed cases and cumulative incidences at 7-days and 14-days across the 18 weeks of the study period.

**Figure 1.**
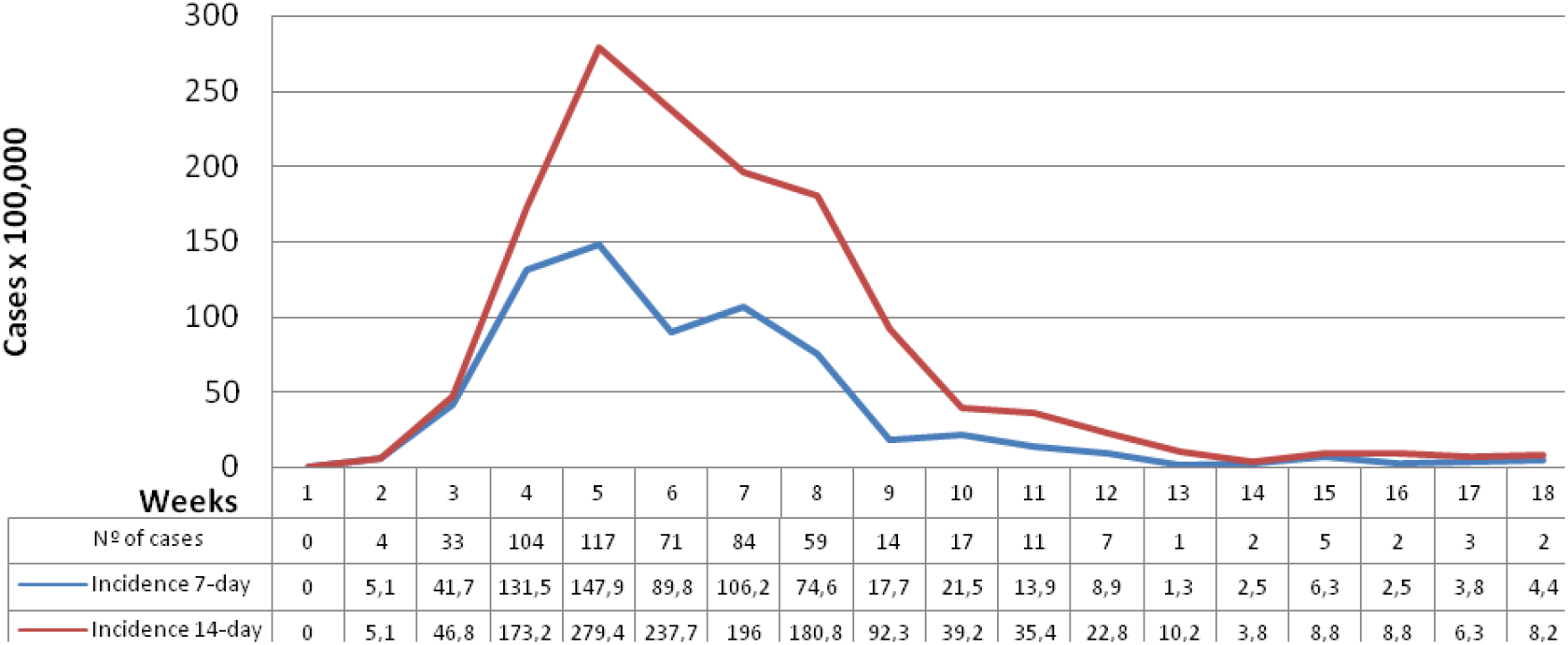
Weekly distribution of COVID19-confirmed cases accross study period (March 1st to June 30th, 2020).

Mean IR across study period was 39.5 (95% CI: 36.1-43.2) cases per 100,000 persons-week (24.1 in 50-64 years vs 30.6 in 65-79 years vs 127.0 in 80 years or older; 36.4 in men vs 42.3 in women; 25.8 in community-dwelling vs 897.2 in nursing-home residents).

By underlying conditions, maximum IRs (per 100,000 persons-week) emerged among those persons with neurological diseases (206.0) followed by atrial fibrillation (109.4), chronic renal failure (85.9), chronic heart disease (66.9), inflammatory bowel disease (66.0), chronic respiratory disease (64.6), cancer (59.6), diabetes (55.4), and hypertension (48.5). Lower IRs were observed among persons with rheumatic diseases (47.0), liver disease (44.0), hypercholesterolemia (38.6), obesity (36.3), alcoholism (35.7) and smokers (18.7).

According to baseline chronic medications, maximum IRs (per 100,000p-w) appeared among those receiving diuretics (97.0), oral anticoagulants (87.3), insulin (83.1), and inhaled-respiratory therapy (72.1) (Table 1).

**Table 1.** Incidence of laboratory-confirmed COVID-19 cases according to baseline demographic and clinical characteristics in the total study cohort (N=79,083). Tarragona region, 01/03/2020-30/06/2020.

| CHARACTERISTIC | Study population<br>(N=79,083)<br>n (%) | LABORATORY-CONFIRMED COVID-19 CASES (n=536) |  |  |
| --- | --- | --- | --- | --- |
|  |  | Univariate analysis<br>n (%) p-value | Time follow-up<br>(persons-week) | Incidence rate |
|  |  |  |  | IR (95% CI) |
| Socio-demographic |  |  |  |  |
| Age: 50-64 years | 42,684 (54.0) | 177 (33.0) <0.001 | 734,123 | 24.1 (20.8-28.0) |
| 65-79 years | 26,013 (32.9) | 137 (25.6) | 447,474 | 30.6 (25.8-36.2) |
| ≥80 years | 10,386 (13.1) | 222 (41.4) | 174,761 | 127.0 (110.2-146.2) |
| Sex Men | 37,626 (47.6) | 235 (43.8) 0.082 | 645,470 | 36.4 (32.1-41.3) |
| Women | 41,457 (52.4) | 301 (56.2) | 710,888 | 42.3 (37.7-47.4) |
| Community-dwelling | 77,669 (98.2) | 345 (64.4) <0.001 | 1,335,069 | 25.8 (23.2-28.7) |
| Nursing-home residence | 1414 (1.8) | 191 (35.6) | 21,289 | 897.2 (778.8-1032.7) |
| Comorbidities |  |  |  |  |
| Neurological disease | 2317 (2.9) | 79 (14.7) <0.001 | 38,344 | 206.0 (164.4-257.5) |
| Renal disease | 4476 (5.7) | 65 (12.1) <0.001 | 75,629 | 85.9 (67.4-109.1) |
| Cancer | 6630 (8.4) | 67 (12.5) 0.001 | 112,481 | 59.6 (46.8-75.7) |
| Rheumatic disease | 872 (1.1) | 7 (1.3) 0.651 | 14,897 | 47.0 (18.8-96.8) |
| Inflammatory bowel disease | 532 (0.7) | 6 (1.1) 0.204 | 9097 | 66.0 (24.2-143.9) |
| Respiratory disease | 7272 (9.2) | 80 (14.9) <0.001 | 123,794 | 64.6 (51.6-80.8) |
| Cardiac disease | 13,435 (17.0) | 153 (28.5) <0.001 | 228,716 | 66.9 (57.1-78.3) |
| Atrial fibrillation | 3786 (4.8) | 70 (13.1) <0.001 | 63,962 | 109.4 (85.9-138.9) |
| Liver disease | 1465 (1.9) | 11 (2.1) 0.731 | 24,998 | 44.0 (22.0-78.8) |
| Diabetes | 13,317 (16.8) | 126 (23.5) <0.001 | 227,640 | 55.4 (46.1-66.5) |
| Hypertension | 34,945 (44.2) | 290 (54.1) <0.001 | 597,890 | 48.5 (43.3-54.4) |
| Hypercholesterolemia | 27,314 (34.5) | 181 (33.8) 0.707 | 468,535 | 38.6 (33.3-44.8) |
| Obesity | 21,678 (27.4) | 135 (25.2) 0.247 | 372,086 | 36.3 (30.6-43.0) |
| Smoking | 12,750 (16.1) | 41 (7.6) <0.001 | 219,518 | 18.7 (13.4-25.4) |
| Alcoholism | 1796 (2.3) | 11 (2.1) 0.733 | 30,774 | 35.7 (17.8-63.9) |
| Chronic medication use |  |  |  |  |
| Diuretics | 8481 (10.7) | 139 (25.9) <0.001 | 143,357 | 97.0 (81.9-114.8) |
| Beta blockers | 9571 (12.1) | 87 (16.2) 0.003 | 163,450 | 53.2 (43.0-66.0) |
| ACEIs | 16,419 (20.8) | 114 (21.3) 0.772 | 281,514 | 40.5 (33.7-48.6) |
| ARBs | 8869 (11.2) | 54 (10.1) 0.401 | 152,376 | 35.4 (26.3-46.7) |
| Calcium channel blockers | 6490 (8.2) | 64 (11.9) 0.002 | 111,107 | 57.6 (44.4-74.9) |
| Statins | 16,134 (20.4) | 91 (17.0) 0.048 | 277,237 | 32.8 (26.5-40.7) |
| Oral anticoagulants | 3912 (4.9) | 58 (10.8) <0.001 | 66,424 | 87.3 (67.2-113.5) |
| Antiplatelet drugs | 9154 (11.6) | 108 (20.1) <0.001 | 155,957 | 69.2 (56.6-84.4) |
| Insulin | 3042 (3.8) | 43 (8.0) <0.001 | 51,741 | 83.1 (60.6-111.4) |
| Oral anti-diabetic drugs | 10,585 (13.4) | 85 (15.9) 0.091 | 181,452 | 46.8 (37.9-58.0) |
| Inhaled respiratory drugs | 6293 (8.0) | 77 (14.4) <0.001 | 106,857 | 72.1 (57.5-90.1) |
| Antineoplastic agents | 1614 (2.0) | 12 (2.2) 0.745 | 27,691 | 43.3 (22.4-75.8) |
| Systemic corticosteroids | 1252 (1.6) | 10 (1.9) 0.599 | 21,030 | 47.6 (22.8-87.6) |
| NSAIDs | 4321 (5.5) | 21 (3.9) 0.114 | 74,152 | 28.3 (17.5-43.3) |
| Chloroquine | 168 (0.2) | 1 (0.2) 0.896 | 2867 | 34.9 (0.9-194.4) |
| Antihistamines | 3264 (4.1) | 15 (2.8) 0.121 | 56,243 | 26.7 (15.0-44.1) |
| Proton-Pump Inhibitors | 17,931 (22.7) | 183 (34.1) <0.001 | 305,566 | 59.9 (51.6-69.5) |
| Benzodiazepines | 13,046 (16.5) | 126 (23.5) <0.001 | 222,537 | 56.6 (47.1-67.9) |
| Vaccination's history |  |  |  |  |
| Flu vaccine in prior autumn | 22,606 (28.6) | 258 (48.1) <0.001 | 385,668 | 66.9 (59.0-75.9) |
| PPV23 | 26,183 (33.1) | 272 (50.7) <0.001 | 447,270 | 60.8 (53.6-68.9) |
| PCV13 | 1139 (1.4) | 18 (3.4) <0.001 | 19,272 | 93.4 (55.4-147.6) |
| Tetanus | 50,799 (64.2) | 344 (64.2) 0.978 | 871,204 | 39.5 (35.5-43.9) |

NOTE: p-values in univariate analysis were calculated by chi-squared, or Fisher's test as appropriate, comparing percentages in the study population vs in COVID-19 cases; IR denotes incidence rates per 100,000 persons-week; CIs denotes confidence intervals for incidence rates and were calculated assuming a Poisson distribution for uncommon events.

Table 2 shows unadjusted, age&sex-adjusted and multivariable-adjusted analyses evaluating risk of suffering laboratory-confirmed COVID19 in the whole study cohort. In the unadjusted analysis, many previous comorbidities and medications were associated with increased risk. However, after multivariable-adjustment, only age (HR: 1.01; 95% CI: 1.00-1.02; p=0.051), nursing-home residence (HR: 20.19; 95% CI: 15.98-25.51; p<0.001), neurological disease (HR: 1.35; 95% CI: 1.03-1.77; p=0.029), taking diuretics (HR: 1.39; 95% CI: 1.10-1.75; p=0.006), antiplatelet drugs (HR: 1.36; 95% CI: 1.05-1.76; p=0.021), benzodiazepines (HR: 1.24; 95% CI: 1.00-1.53; p=0.047) and history of PCV13 vaccination (HR: 1.74; 95% CI: 1.06-2.85; p=0.028) appeared significantly associated with increased risk. Oppositely, smoking (HR: 0.57; 95%CI: 0.41-0.80; p=0.001), taking angiotensin-converting enzyme (ACE) inhibitors (HR: 0.78; 95% CI: 0.61-1.00; p=0.048), angiotensin II receptor blockers -ARBs-(HR: 0.70; 95%CI: 0.51-0.96; p=0.027) and statins (HR: 0.75; 95% CI: 0.58-0.96; p=0.025) appeared associated with a reduced risk.

**Table 2.** Cox regression analyses assessing unadjusted, age & sex adjusted and multivariable-adjusted risks to suffer laboratory-confirmed COVID-19 in the total study cohort (N=79,083). Tarragona region (Southern Catalonia, Spain), from 01/03/2020 to 30/06/2020.

| CHARACTERISTIC | LABORATORY-CONFIRMED COVID-19 CASES (n=536) |  |  |
| --- | --- | --- | --- |
|  | Unadjusted<br>HR (95% CI) p-value | Age & sex adjusted<br>HR (95% CI) p-value | Multivariable-adjusted<br>HR (95% CI) p-value |
| **Socio-demographic** |  |  |  |
| **Age (continuous years)** | 1.06 (1.05-1.07) <0.001 | 1.06 (1.05-1.07) <0.001 | 1.01 (1.00-1.02) 0.051 |
| **Sex: women** | 1.16 (0.98-1.38) 0.083 | 1.00 (0.84-1.19) 0.992 | 0.89 (0.74-1.07) 0.214 |
| **Nursing-home residence** | 33.18 (27.80-39.59) <0.001 | 23.80 (19.18-29.53) <0.001 | 20.19 (15.98-25.51) <0.001 |
| **Comorbidities** |  |  |  |
| **Neurological disease** | 5.86 (4.62-7.44) <0.001 | 3.01 (2.33-3.88) <0.001 | 1.35 (1.03-1.77) 0.029 |
| **Renal disease** | 2.33 (1.80-3.02) <0.001 | 1.09 (0.83-1.44) 0.518 | 1.01 (0.76-1.34) 0.959 |
| **Cancer** | 1.57 (1.22-2.03) 0.001 | 1.16 (0.90-1.50) 0.264 | 1.18 (0.91-1.55) 0.216 |
| **Rheumatic disease** | 1.19 (0.57-2.51) 0.646 | 1.07 (0.51-2.25) 0.866 | 1.29 (0.59-2.84) 0.519 |
| **Respiratory disease** | 1.74 (1.38-2.21) <0.001 | 1.36 (1.07-1.73) 0.012 | 1.16 (0.83-1.62) 0.377 |
| **Cardiac disease** | 1.97 (1.63-2.37) <0.001 | 1.21 (1.00-1.48) 0.054 | 0.99 (0.79-1.25) 0.950 |
| **Atrial fibrillation** | 3.02 (2.35-3.89) <0.001 | 1.57 (1.20-2.04) 0.001 | 1.25 (0.83-1.88) 0.281 |
| **Liver disease** | 1.12 (0.61-2.03) 0.721 | 1.26 (0.69-2.28) 0.453 | 1.13 (0.62-2.06) 0.699 |
| **Diabetes** | 1.52 (1.25-1.86) <0.001 | 1.13 (0.92-1.38) 0.243 | 1.09 (0.76-1.55) 0.644 |
| **Hypertension** | 1.49 (1.26-1.77) <0.001 | 0.83 (0.69-1.00) 0.050 | 0.97 (0.77-1.23) 0.823 |
| **Obesity** | 0.89 (0.73-1.08) 0.245 | 0.81 (0.66-0.98) 0.032 | 0.90 (0.73-1.11) 0.328 |
| **Smoking** | 0.43 (0.31-0.59) <0.001 | 0.69 (0.49-0.95) 0.024 | 0.57 (0.41-0.80) 0.001 |
| **Alcoholism** | 0.90 (0.50-1.64) 0.735 | 1.16 (0.64-2.13) 0.623 | 0.96 (0.52-1.77) 0.896 |
| **Chronic medication use** |  |  |  |
| **Diuretics** | 2.95 (2.43-3.58) <0.001 | 1.71 (1.39-2.10) <0.001 | 1.39 (1.10-1.75) 0.006 |
| **Beta blockers** | 1.41 (1.12-1.78) 0.003 | 1.01 (0.80-1.28) 0.920 | 0.94 (0.73-1.22) 0.664 |
| **ACEIs** | 1.03 (0.84-1.27) 0.769 | 0.75 (0.61-0.92) 0.007 | 0.78 (0.61-1.00) 0.048 |
| **ARBs** | 0.89 (0.67-1.17) 0.399 | 0.62 (0.47-0.82) 0.001 | 0.70 (0.51-0.96) 0.027 |
| **Calcium channel blockers** | 1.52 (1.17-1.97) 0.002 | 1.04 (0.80-1.35) 0.793 | 1.19 (0.90-1.58) 0.225 |
| **Statins** | 0.80 (0.64-1.00) 0.049 | 0.60 (0.48-0.75) <0.001 | 0.75 (0.58-0.96) 0.025 |
| **Oral anticoagulants** | 2.35 (1.79-3.09) <0.001 | 1.29 (0.98-1.71) 0.074 | 1.18 (0.76-1.84) 0.456 |
| **Antiplatelet drugs** | 1.94 (1.57-2.39) <0.001 | 1.21 (0.97-1.51) 0.085 | 1.36 (1.05-1.76) 0.021 |
| **Insulin** | 2.20 (1.61-3.00) <0.001 | 1.64 (1.20-2.25) 0.002 | 1.24 (0.85-1.81) 0.261 |
| **Oral anti-diabetic drugs** | 1.22 (0.97-1.54) 0.091 | 0.94 (0.74-1.18) 0.584 | 1.02 (0.70-1.48) 0.922 |
| **Inhaled respiratory drugs** | 1.96 (1.54-2.49) <0.001 | 1.43 (1.12-1.83) 0.004 | 1.23 (0.87-1.72) 0.242 |
| **Antineoplastic agents** | 1.10 (0.62-1.95) 0.746 | 1.04 (0.59-1.84) 0.899 | 0.94 (0.51-1.73) 0.841 |
| **Systemic corticosteroids** | 1.20 (0.64-2.24) 0.574 | 0.88 (0.47-1.65) 0.687 | 0.80 (0.42-1.53) 0.499 |
| **NSAIDs** | 0.71 (0.46-1.09) 0.118 | 0.84 (0.54-1.30) 0.433 | 1.13 (0.72-1.76) 0.603 |
| **Antihistamines** | 0.67 (0.40-1.12) 0.122 | 0.68 (0.41-1.14) 0.144 | 0.73 (0.43-1.22) 0.231 |
| **Proton-Pump Inhibitors** | 1.78 (1.49-2.13) <0.001 | 1.11 (0.92-1.34) 0.276 | 0.92 (0.74-1.15) 0.464 |
| **Benzodiazepines** | 1.56 (1.28-1.91) <0.001 | 1.19 (0.97-1.46) 0.092 | 1.24 (1.00-1.53) 0.047 |
| **Vaccination's history** |  |  |  |
| **Flu vaccine (prior autumn)** | 2.33 (1.97-2.76) <0.001 | 1.27 (1.05-1.54) 0.013 | 1.00 (0.80-1.25) 0.987 |
| **PPV23** | 2.09 (1.77-2.48) <0.001 | 0.85 (0.69-1.04) 0.106 | 1.05 (0.82-1.35) 0.693 |
| **PCV13** | 2.40 (1.50-3.84) <0.001 | 1.79 (1.12-2.87) 0.015 | 1.74 (1.06-2.85) 0.028 |
| **Tetanus** | 1.00 (0.84-1.19) 0.983 | 0.73 (0.61-0.87) <0.001 | 0.84 (0.68-1.03) 0.091 |

NOTE: HRs denotes Hazard ratios, and were calculated for those who had the condition as compared with those who had not the condition. In multivariable-adjusted analysis the HRs were adjusted for age (continuous years), sex, residence, comorbidities/underlying conditions and chronic medication use. CIs denote confidence intervals.

Table 3 (incidence rates) and Table 4 (unadjusted and adjusted risks) show subgroup analyses restricted to community-dwelling persons (N=77,669). In the multivariable analysis focused on these individuals, chronic renal disease (HR: 1.50; 95% CI: 1.02-2.21; p=0.038), cancer (HR: 1.45; 95% CI: 1.05-2.01; p=0.023), cardiac disease (HR: 1.32; 95% CI: 0.98-1.79; p=0.070), taking diuretics (HR: 1.54; 95% CI: 1.11-2.13; p=0.010) and benzodiazepines (HR: 1.28; 95% CI: 0.97-1.68; p=0.078)were associated (significantly or near significantly) with increased risk. Oppositely, smoking (HR: 0.51; 95% CI: 0.35-0.74; p<0.001), receiving ACE-inhibitors (HR: 0.63;95% CI: 0.45-0.89; p=0.009), statins (HR: 0.66; 95% CI: 0.49-0.90; p=0.009) and influenza vaccination in prior autumn (HR: 0.68; 95% CI: 0.51-0.92; p=0.012) were associated with decreased risk.

**Table 3.**
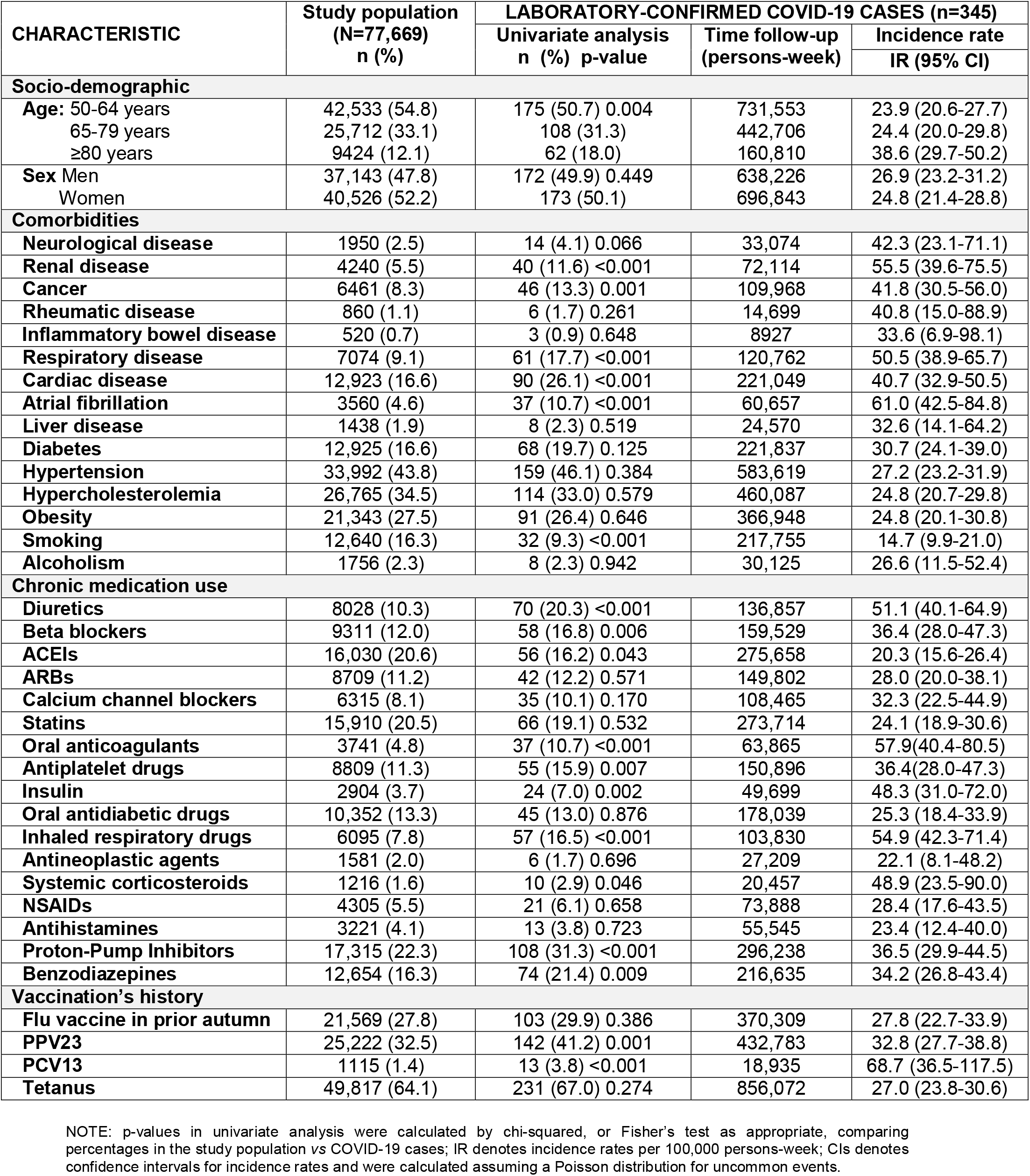
Incidence of laboratory-confirmed COVID-19 cases according to baseline demographic and clinical characteristics (comorbidities/medications) in community-dwelling individuals (N=77,669). Tarragona region (Southern Catalonia, Spain), 01/03/2020-30/06/2020.

**Table 4.**
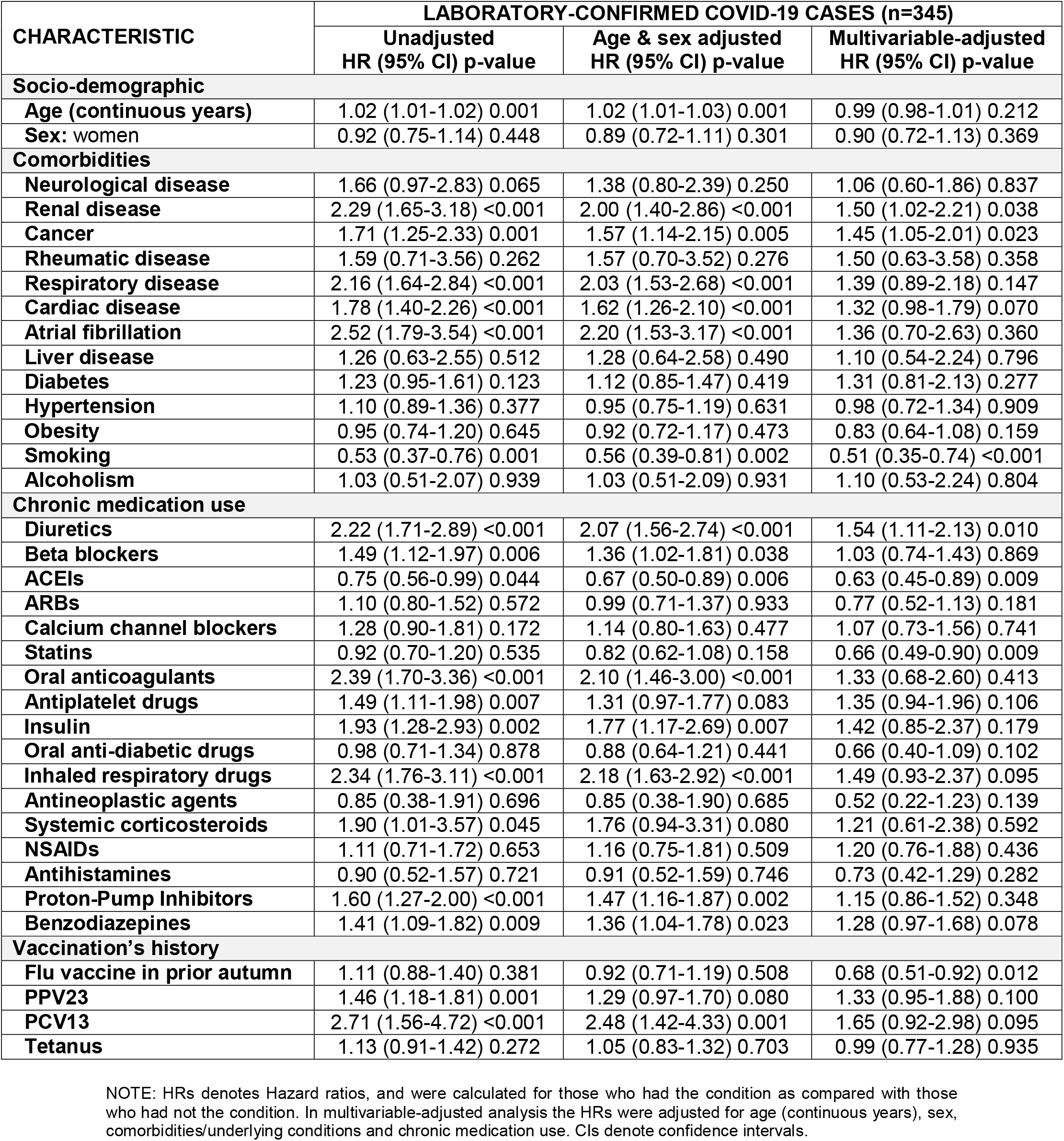
Cox regression analyses assessing unadjusted, age & sex adjusted and multivariable-adjusted risks to suffer laboratory-confirmed COVID-19 in community-dwelling individuals (N=77,669). Tarragona region (Southern Catalonia, Spain) from 01/03/2020 to 30/06/2020.

Table 5 shows subgroup analyses restricted to nursing-home residents (N=1414). In the multivariable analysis, increasing age (HR: 1.03; 95% CI: 1.01-1.05; p=0.001), oral anti-diabetic drugs (HR: 1.79; 95% CI: 1.04-3.08; p=0.037), antineoplastic agents (HR: 2.51; 95% CI: 1.05-6.01; p=0.039) and influenza vaccination in prior autumn (HR: 1.56; 95% CI: 1.07-2.29; p=0.022) were associated with increased risk. In contrast, ARBs (HR: 0.47; 95% CI: 0.25-0.87; p=0.017) and history of tetanus vaccination (HR: 0.63; 95% CI: 0.44-0.91; p=0.013) were associated with decreased risk.

**Table 5.** Univariate and multivariate analyses assessing susceptibility/risk to suffer laboratory-confirmed Covid-19 among nursing-home residents (N=1,414). Tarragona region (Southern Catalonia, Spain), 01/03/2020-30/06/2020.

| CHARACTERISTIC | Study population<br>(N=1414)<br>n (%) | LABORATORY-CONFIRMED Covid-19 CASES (n=191) |  |
| --- | --- | --- | --- |
|  |  | Univariate analysis<br>n (%) p value | Multivariable analysis<br>HR (95% CI) p value |
| Socio-demographic |  |  |  |
| Age: 50-64 years | 151 (10.7) | 2 (1.0) <0.001 | Age, continuous years:<br>1.03 (1.01-1.05) 0.001 |
| 65-79 years | 301 (21.3) | 29 (15.2) |  |
| ≥80 years | 962 (68.0) | 160 (83.8) |  |
| Sex: Men | 483 (34.2) | 63 (33.9) 0.713 | 1.00 (reference) 0.001 |
| Women | 931 (65.8) | 128 (66.1) | 0.78 (0.56-1.09) 0.140 |
| Comorbidities |  |  |  |
| Neurological disease | 367 (26.0) | 65 (34.0) 0.006 | 1.28 (0.93-1.75) 0.127 |
| Renal disease | 236 (16.7) | 25 (13.1) 0.151 | 0.69 (0.44-1.08) 0.106 |
| Cancer | 169 (12.0) | 21 (11.0) 0.661 | 0.83 (0.51-1.35) 0.462 |
| Rheumatic disease | 12 (0.8) | 1 (0.5) 0.598 | 0.73 (0.10-5.42) 0.762 |
| Respiratory disease | 198 (14.0) | 19 (9.9) 0.082 | 0.75 (0.43-1.32) 0.315 |
| Cardiac disease | 512 (36.2) | 63 (33.0) 0.319 | 0.80 (0.57-1.12) 0.188 |
| Atrial fibrillation | 226 (16.0) | 33 (17.3) 0.600 | 1.23 (0.72-2.08) 0.447 |
| Liver disease | 27 (1.9) | 3 (16) 0.713 | 0.83 (0.26-2.63) 0.744 |
| Diabetes | 392 (27.7) | 58 (30.4) 0.380 | 1.02 (0.61-1.68) 0.955 |
| Hypertension | 953 (67.4) | 131 (68.6) 0.706 | 0.98 (0.68-1.41) 0.906 |
| Obesity | 335 (23.7) | 44 (23.0) 0.819 | 1.13 (0.79-1.61) 0.520 |
| Smoking | 110 (7.8) | 9 (4.7) 0.089 | 0.98 (0.47-2.02) 0.952 |
| Alcoholism | 40 (2.8) | 3 (1.6) 0.259 | 0.59 (0.18-1.92) 0.379 |
| Chronic medication use |  |  |  |
| Diuretics | 453 (32.0) | 69 (36.1) 0.193 | 1.26 (0.91-1.75) 0.162 |
| Beta blockers | 260 (18.4) | 29 (15.2) 0.219 | 0.77 (0.50-1.19) 0.243 |
| ACEIs | 389 (27.5) | 58 (30.4) 0.342 | 0.97 (0.68-1.38) 0.867 |
| ARBs | 160 (11.3) | 12 (6.3) 0.018 | 0.47 (0.25-0.87) 0.017 |
| Calcium channel blockers | 175 (12.4) | 29 (15.2) 0.205 | 1.37 (0.89-2.10) 0.156 |
| Statins | 224 (15.8) | 25 (13.1) 0.263 | 0.92 (0.58-1.48) 0.739 |
| Oral anticoagulants | 171 (12.1) | 21 (11.0) 0.617 | 0.87 (0.46-1.65) 0.675 |
| Antiplatelet drugs | 345 (24.4) | 53 (27.7) 0.246 | 1.35 (0.92-1.99) 0.123 |
| Insulin | 138 (9.8) | 19 (9.9) 0.925 | 0.82 (0.47-1.43) 0.481 |
| Oral anti-diabetic drugs | 233 (16.5) | 40 (20.9) 0.074 | 1.79 (1.04-3.08) 0.037 |
| Inhaled respiratory drugs | 198 (14.0) | 20 (10.5) 0.130 | 0.81 (0.47-1.40) 0.447 |
| Antineoplastic agents | 33 (2.3) | 6 (3.1) 0.427 | 2.51 (1.05-6.01) 0.039 |
| Systemic corticosteroids | 36 (2.5) | 0 (0.0) 0.016 | 0.00 (-) 0.945 |
| NSAIDs | 16 (1.1) | 0 (0.0) 0.112 | 0.00 (-) 0.967 |
| Antihistamines | 43 (3.0) | 2 (1.0) 0.084 | 0.40 (0.10-1.63) 0.199 |
| Proton-Pump Inhibitors | 616 (43.6) | 75 (39.3) 0.198 | 0.74 (0.53-1.04) 0.083 |
| Benzodiazepines | 392 (27.7) | 52 (27.2) 0.869 | 0.97 (0.70-1.36) 0.879 |
| Vaccination's history |  |  |  |
| Flu vaccine in prior autumn | 1037 (73.3) | 155 (81.2) 0.009 | 1.56 (1.07-2.29) 0.022 |
| PPV23 | 961 (68.0) | 130 (68.1) 0.975 | 0.95 (0.65-1.39) 0.778 |
| PCV13 | 24 (1.7) | 5 (2.6) 0.290 | 1.89 (0.73-4.88) 0.187 |
| Tetanus | 982 (69.4) | 113 (59.2) 0.001 | 0.63 (0.44-0.91) 0.013 |

NOTE: p-values in univariate analysis were calculated by chi-squared, or Fisher's test as appropriate, comparing percentages in the study population vs COVID-19 cases; HRs denotes Hazard ratios, and were calculated for those who had the condition as compared with those who had not the condition, and were adjusted for age (continuous years), sex, comorbidities/underlying conditions and chronic medication use. CIs denote confidence intervals.

## DISCUSSION

Population-based clinical data on susceptibility and risk of SARS-COV-2 infection is limited. [9, 18] This large cohort study was aimed to assess population-based incidence of (laboratory-confirmed) COVID19 among middle-aged and older adults with distinct underlying medical conditions in a well characterised geographic area (Tarragona region) during the first wave of the COVID19 epidemic (March-June 2020); we investigated possible relationships between pre-existing medical conditions (including common comorbidities, vaccinations’ history and chronic medication use) with the susceptibility/risk of suffering COVID19.

As main result, we found a mean IR of 39.5 laboratory-confirmed COVID19 cases per 100,000 persons-week thru study period, which is a relatively low incidence as compared to other Spanish or European regions during the same time period. [19,20].

During the study period (17.4 weeks since 1st March until 30th June 2020), approximately 5% of cohort members were PCR tested (with 12.4% positive results). Taking this data into account, and considering that PCR test were scarcely available in the study area for patients with less severe symptoms during the first weeks of the epidemic period, the true incidence of COVID19 was logically underestimated. Nevertheless, considering the relatively low number of presumptive cases (clinical suspicion alone without PCR performed), our data also suggests that the overall number of infected people was considerably lower than initially speculated (as showed by the national seroprevalence study). [21] Assuming this relatively little incidence, we found that increasing age, nursing-home residence, some pre-existing comorbidities (cancer, neurological, renal or cardiac disease and taking certain chronic medications (diuretics, antiplatelet drugs and benzodiazepines) were independent factors related to increased risk of suffering COVID19. In contrast, cohort members receiving statins or ACEIs/ARBs (and, in the case of community-dwelling individuals, those vaccinated against influenza in prior autumn) presented, after multivariable adjustment, a decreased risk.

Of note, in the crude analyses several covariates (comorbidities, medications and vaccinations) appeared significantly associated with increased risk/susceptibility of suffering COVID19. However, for most of them the significance disappeared after age&sex-adjustment and only a few remained independently associated with an increased risk after multivariable adjustments. This highlights the importance of reporting adjusted data and note that differences in adjustment methods may explain, in part, the heterogeneous results (when evaluating the possible association between some pre-existing conditions and risk of COVID19) reported to date. [22] At present, there is not clear evidence about possible clinical predisposing and protecting factors front SARS-COV-2 infection, and uncertainty surrounds this issue. [23]

If we consider demographic characteristics, apart from nursing-home residence that increased twenty-times the adjusted-risk of suffering laboratory-confirmed COVID19, we found that age increased approximately 1-2% for each year the adjusted-risk of suffering COVID19. While COVID19 was more frequent in women, sex did not alter significantly the risk of infection in multivariable analysis.

Considering pre-existing comorbidities, only neurological disease (basically patients with a history of dementia and/or stroke living in nursing-home residences) was associated with increased multivariable-adjusted risk of infection in the analysis of full cohort. Concomitantly, previous cancer, renal disease and cardiac disease emerged significantly associated with increased risk in subgroup analysis restricted to community-dwelling individuals.

In the present study, chronic respiratory disease, diabetes, hypertension and obesity did not emerge independently associated with significant increased risk of suffering laboratory-confirmed COVID19 after multivariable-adjusted analyses. There is general accordance considering these conditions as major risk factors for poor prognosis in hospitalised COVID19 patients, but there is few data assessing their role in predisposing for SARS-COV-2 infection. [24]

Surprisingly, smokers presented the lowest specific incidence rate of COVID19 in the present report, and smoking was associated with a statistically significant reduced risk of COVID19, both in multivariable analyses evaluating the total study cohort and in the restricted to community-dwelling individuals. Notably, active smoking has been linked with decreased odds of a positive result SARS-CoV-2-test in other studies. [8] Opposite findings about poor prognosis among smokers with COVID19 have been reported. [25,26]

Considering chronic medication use, receiving diuretics, antiplatelet drugs and benzodiazepines appeared significantly associated with increased risk of suffering COVID19. In contrast, taking statins and ACEIs or ARBs emerged associated with decreased risk. At onset of COVID19 pandemic, after discovering that the host receptor for SARS-CoV-2 cell entry was the angiotensin-converting enzyme 2 (ACE2), it was feared that receiving ACEIs/ARBs could facilitate SARS-COV-2 infection and predispose to suffer severe COVID-19. However, most recent studies have concluded that there is no clinical or experimental evidence supporting that ACEIs or ARBs augment the susceptibility to/severity of COVID-19 at present. Indeed, ACEIs/ARBs have been associated with lower incidence and/or improved outcomes in patients with lower respiratory tract infections, and with lower risk of all-cause mortality in COVID19 hospitalised patients. Our data also support this conclusion.

Considering statins, while first studies reported opposite results, our data showing a decreased risk of laboratory-confirmed COVID19 among people taking these drugs are in accordance with conclusions of a recent meta-analysis on this concern. [28] Anyway, caution is recommended until further evidence emerges surrounding statin use in COVID19 patients.

Regarding other chronic medications that appeared associated with increased risk in the current study (i.e., diuretics, antiplatelet drugs and benzodiazepines), we have not a clear interpretation for this data. It is possibly due to an unresolved residual confounding, linked to unmeasured health-care-related factors or insufficient age-adjustment, but we also note that some of them (e.g., benzodiazepines) have already been reported associated with greater risk of pneumonia (common clinical manifestation in COVID19 patients).

Antihistamines’ use was not associated with any statistically significant change in the risk of suffering COVID-19 in the present study. We note that antihistamines’ use was associated (marginally significative) with a lower risk of COVID19 in an earlier report, [12] but we also remark that it could be due to an alpha error considering that this final report has greater statistical power (since include more persons-time follow-up and greater number of events analysed).

Similar to data observed in earlier analyses, we found that influenza vaccination in prior autumn was significantly associated with a reduction risk of suffering COVID19 among community-dwelling individuals. [10] Conversely, it was associated with a greater risk in the subgroup of nursing-home residents (where a significant protective association appeared for those who had received tetanus vaccination). While these associations could be due to residual confounding related with unmeasured life-style or healthcare factors, further investigations are needed exploring a possible immunity-related mechanism explanation. [29,30]

Major strengths in this study were the representativity (due to large size) of the study cohort, that included almost 80,000 people, which represents about 75% of the entire population over 50 years in the area, [13] and the use of survival analysis to precisely asses susceptibility and risk of suffering a SARS-COV-2 infection adjusting by important potential confounders (as socio-demographic, and medical underlying conditions clinically relevant)

Major limitations were related to its partially retrospective design and poor access to PCR tests at the beginning of the epidemic period in our setting, especially considering that the most specific diagnosis for COVID19 is a positive PCR test. Moreover its reliability depends on the correct collection of nasopharyngeal swabs, moment of collection, sensitivity of tests and guidelines for testing followed throughout the study period. Due to above mentioned scarce availability of PCR tests they were mainly performed in hospitalised/severe patients and in nursing homes (after some outbreaks were detected). Thereby, patients with mild symptoms (many of them without PCR done) have been logically underestimated in this study.

Even though we did subgroup analyses (community-dwelling/nursing-home) and multivariable-adjustments, residual confounding when estimating risks (common in observational studies), tied to non-included variables such as socio-economical, labor-, lifestyle- or health care-related factors, may not be discarded.

Despite the large size of the cohort, there were relatively few events (536 laboratory-confirmed COVID19 cases), which limits statistical power. This is especially relevant in the subgroup of nursing-home residents where, even if great incidence occurred (200 cases per 100,000 persons-week), only 191 confirmed COVID19 cases were detected, fact that largely affects the precision of estimates (confidence intervals very wide).

Notwithstanding all mentioned limitations, authors may emphasise the added value that provide adjustments (age&sex and multivariable), not present in many other studies. Their importance is clearly evident if we compare, for example in Table 3 and 5, crude with adjusted results.

Concerning external validity, we must consider that the study was conducted in a single geographic area; thus, specific incidence data cannot be directly extrapolated to other regions with distinct epidemic conditions.

Nevertheless, we also note that the adjusted-risks estimated here may be helpful to better characterize the risk profile (socio-demographic, pre-existing conditions/comorbidities, chronic medication use) for suffering COVID19 among middle-aged and older adults living in a setting with low/moderate intensity of COVID19, as in the present study.

## CONCLUSIONS

In summary, in this large population-based study that followed a cohort of 79,083 individuals ≥50 years old between March-June 2020 in Tarragona area, the incidence of laboratory-confirmed SARS-COV-2 infection was relatively low (39.5 cases per 100,000 persons-week) as compared with that of other Spanish regions during the first wave of COVID19 pandemic.

In this epidemic context (low incidence), our data support that increasing age, nursing-home residence, pre-existing cancer, neurological, renal and cardiac disease are independent major predisposing conditions to suffer COVID19 among middle-aged and older adults. Patients receiving diuretics, antiplatelet drugs and benzodiazepines were also at increased risk. Oppositely, patients taking statins, rennin-angiotensin-aldosterone system inhibitors and those community-dwelling individuals that received influenza vaccination in prior autumn presented decreased risk.

## Data Availability

Data are available under appropriate request.

## ACKNOWLEDGEMENTS

This study was supported by a grant from the Instituto de Salud Carlos III of the Spanish Health Ministry (file COV20/00852; call for the SARS-COV-2/COVID-19 disease, RDL 8/2020, March 17, 2020).

## CONTRIBUTORS

ESG and AVC wrote the manuscript; CDC, CTF, FGB, and IHG obtained data; ESG and AVR assessed outcomes and did statistical analyses; AVR edited the manuscript; OOG and FML revised the final version; AVC coordinated the study.

